# Deep neural frameworks improve the accuracy of general practitioners in the classification of pigmented skin lesions

**DOI:** 10.1101/2020.05.03.20072454

**Authors:** Maximiliano Lucius, Jorge De All, José Antonio De All, Martín Belvisi, Luciana Radizza, Marisa Lanfranconi, Victoria Lorenzatti, Carlos M. Galmarini

## Abstract

Artificial intelligence can be a key tool in the context of assisting in the diagnosis of dermatological conditions, particularly when performed by general practitioners with limited or no access to high resolution optical equipment. This study evaluates the performance of deep convolutional neural networks (DNNs) in the classification of seven pigmented skin lesions. Additionally, it assesses the improvement ratio in the classification performance when utilized by general practitioners. Open-source skin images were downloaded from the ISIC archive. Different DNNs (n=8) were trained based on a random dataset constituted by 8,015 images. A test set of 2,003 images has been used to assess the classifiers performance at low (300 × 224 RGB) and high (600 × 450 RGB) image resolution and aggregated clinical data (age, sex and lesion localization). We have also organized two different contests to compare the DNNs performance to that of general practitioners by means of unassisted image observation. Both at low and high image resolution, the DNNs framework being trained differentiated dermatological images with appreciable performance. In all cases, accuracy has been improved when adding clinical data to the framework. Finally, the lowest accurate DNN outperformed general practitioners. Physician’s accuracy was statistically improved when allowed to use the output of this algorithmic framework as guidance. DNNS are proven to be high performers as skin lesion classifiers. The aim is to include these AI tools in the context of general practitioners whilst improving their diagnosis accuracy in a routine clinical scenario when or where the use of high-resolution equipment is not accessible.

## 1. Introduction

Diagnosis in dermatology is largely based on visual inspection of a lesion on the suspicious skin area. Therefore, diagnostic ability and accuracy depends greatly on the experience and training of dermatologists or general practitioners, in areas where dermatological services are not readily available [1]. When dermatologists get no access to additional technical support, they have an approximately 65%-70% accuracy rate in melanoma diagnosis [2-4]. If the lesion is suspicious, the visual inspection is supplemented with different diagnostic tools (e.g. dermoscopy, confocal microscopy or optical coherence tomography) providing the ability to explore the skin *in vivo*, in depth and at a higher resolution [5, 6]. However, access to these instruments remains limited due to time, logistical and cost concerns. Even when this technical support is feasible, dermatologists rarely achieve average rates greater than 85% [7, 8]. The situation is even worse if we consider that there is a shortage of dermatologists whilst diagnostic accuracy of non-expert clinicians is sensibly below than what is observed with dermatologists, reaching estimate rates between 20 and 40% [9-13]. Thus, new diagnostic tools assisting dermatologists or general practitioners to accurately diagnose skin lesions should be developed, evaluated and optimized.

Artificial intelligence (AI) is a computer science that involves creating sequences of data-related instructions that aim to reproduce human cognition [14]. The use of AI to assist physicians has been applied to various medical fields. In dermatology, image recognition using a set of algorithms called deep neural networks (DNNs) have proven to be of significant aid to physicians in the diagnosis of pigmented skin lesions. These algorithms achieve accuracies comparable to those of dermatologists [1, 15-18]. In addition, Hekler et al. demonstrated that the combination of human and artificial intelligence is superior to the individual results of dermatologists or DNNs in isolation [19]. Similar results were observed in the case of non-pigmented skin lesions such as acne, rosacea, psoriasis, atopic dermatitis or impetigo. Thus, these technologies show tremendous promise to improve skin lesion diagnosis and may extend screening far beyond the clinical setting. However, many aspects of their use have yet to be elucidated and improved.

This study has aimed to compare and improve the precision of different digital image-based DNNs models to classify seven different types of conditions representing more than 90% of the pigmented skin lesions. For this purpose, we (i) compare the accuracy of 8 different DNNs in different training conditions such as input of low and high image resolution and with or without clinical data; (ii) evaluate DNNs performance on synthetic images generated with an infoGAN [20]; (iii) compare DNNs accuracy against non-dermatologist general practitioners; (iv) assess if these physicians improved their classification performance when using the framework as an assisting tool.

## 2. Material and Methods

### 2.1 Pigmented skin image dataset

This study used images from the anonymous and annotated HAM10000 dataset publicly available through the International Skin Imaging Collaboration (ISIC) archive [21]. All downloaded images were selected using a random generator from the set of available images in the ISIC archive. We stochastically split the master set of 10,015 dermoscopic images into training (n=8,313; 83%) and test (N=1,702; 17%) datasets that were completely disjoint. Images included a representative collection of all-important diagnostic categories acroos the seven different types of pigmented lesions as detailed in Tschandl et al [21]. Those included melanocytic nevus, vascular skin lesions (including cherry angiomas, angiokeratomas, pyogenic granulomas and hemorrhages), benign keratoses (including seborrheic keratoses, solar lentigo and lichen-planus-like kertoses), dermatofibroma, intraepithelial carcinoma (including actinic keratoses and Bowen’s disease), basal cell carcinoma and melanoma. Examples of images of each lesion type are depicted in Figure 1. The final composition of each dataset is shown in Table 1.

**Figure 1.**
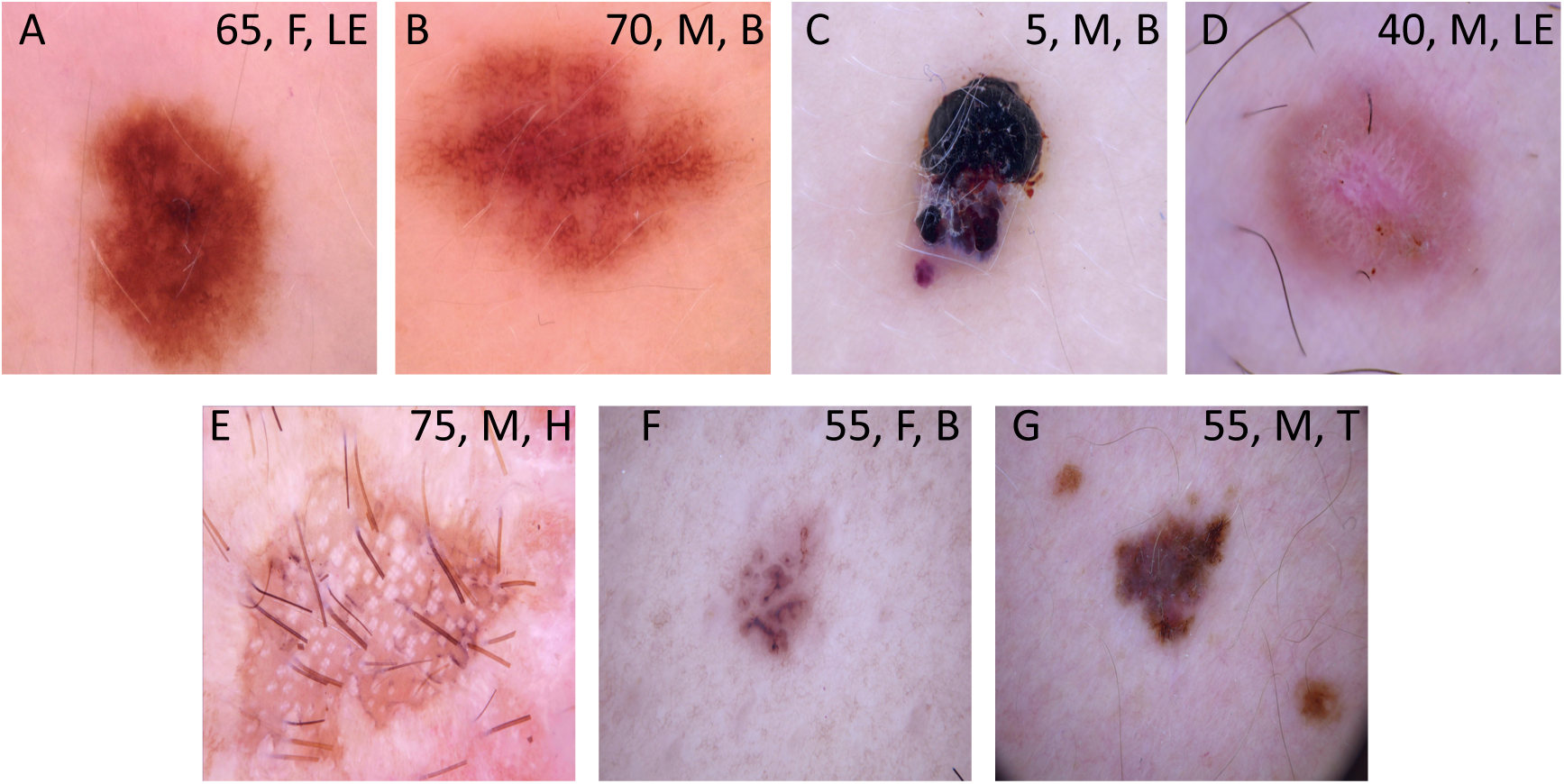
Examples of images downloaded from the HAM10000 dataset. These images are publicly available through the International Skin Imaging Collaboration (ISIC) archive and represent more than 95% of all pigmented lesions encountered during clinical practice (Tschandl P 2018). (A) Melanocytic nevus; (B) Bening keratosis; (C) Vascular lesion; (D) Dermatofibroma; (E) Intraepithelial carcinoma; (F) Basal cell carcinoma; (G) Melanoma. Legends inside each image represents clinical data such as age, sex and localization associated to the image. F: female; M: male; LE: lower extremity; B: back; H: Hand; T: trunk.

**Table 1.** Description of the training and test datasets

| Class | Training set (n) | Test set (n) |
| --- | --- | --- |
| Melanocytic nevi | 5,565 | 1,140 |
| Bening keratosis<sup>1</sup> | 912 | 186 |
| Vascular lesions<sup>2</sup> | 118 | 24 |
| Dermatofibroma | 96 | 20 |
| Intraepithelial carcinoma<sup>3</sup> | 271 | 56 |
| Basal cell carcinoma | 427 | 87 |
| Melanoma | 924 | 189 |
| Total | 8,313 | 1,702 |

<sup>1</sup> includes seborrheic keratoses, solar lentigo and lichen-planus like keratoses
<sup>2</sup> includes cherry angiomas, angiokeratomas, pyogenic granulomas and hemorrhage
<sup>3</sup> includes actinic keratoses and intraepithelial carcinoma (Bowen's disease)

### 2.2 Deep neural networks

We have evaluated eight different DNNs each characterized by a specific architecture. VGG16 and VGG19 contain 16 and 19 convolutional layers respectively, with very small receptive fields, five max-pooling layers of size for carrying out spatial pooling, followed by three fully connected layers, with the final layer as the soft-max layer [22]. Rectification nonlinearity (ReLu) activation is applied to all hidden layers. The model also uses dropout regularization in the fully connected layers. ResNet34 is a 34-layer residual network while ResNet50 and ResNet101 are 50- and 101-layers deep, respectively. The architecture of all these DNNs is similar to the one found in VGG consisting mostly of 3×3 filters but, instead, shortcut connections are inserted resulting into a residual network. SEResNet50 architecture is based on ResNet. A squeeze-and-excitation block is applied at the end of each non-identity branch of residual block [23]. Differently, and instead of increasing its size by adding more or deeper layers, EfficientNetB5 scale up the network width, depth and resolution with a set of fixed scaling coefficients [24]. Finally, MobileNet uses depth-wise separable convolutions which significantly reduces the number of parameters when compared to a network based on standard convolutions and the same depth across the structure in the networks. The framework is 54 layers deep.

To assess both the performance of the algorithm and the enhanced training techniques as accurately as possible, we have retrained each DNN a total of 5 times (folds), and each training run consisted of 90 epochs. Training using the curated image patches took approximately 6 h to complete, 45k iterations on a 4 GeForce GTX 1080 GPU configuration. Training accuracy for curated patches reached maximum accuracy (100%) at around epoch 32, whereas the pretrained model only began to converge around epoch 25. All these DNNs were trained and tested with two different image input size: 300 × 224 RGB and 600 × 450 RGB. The low-resolution images were obtained by cropping, distorting and linear resizing the original high-resolution images. DNNs were also trained and tested without or with the clinical features (sex, age and location of the lesion) associated to every image in the HAM10000 database.

### 2.3 Image preprocessing

When a deep convolutional neural network overfits, it works extremely well on training data but poorly on data it has never seen before. This is especially important in the field of dermatology because of the variability that exists in the images that the neural network will be analyzing. Two steps were taken to reduce overfitting. First, a dropout layer was added and set to 0.5. This results in 50% of the neurons to be randomly turned off during the training process and therefore reduce the likelihood of overfitting. The second step taken to reduce overfitting was to use data augmentation. In data augmentation, the images are modified to account for some of the variability that exists in image taking. To account for grid location, size of the dermatological manifestation and the angle of the image, the training images fed into the model were altered using rotation, zoom, shear, and horizontal and vertical flipping randomly or following optical parameters from the different types of phone cameras. The model was run once without data augmentation and but with dropout and once with data augmentation including drop out in both instances.

### 2.4 Generation of synthetic pigmented skin lesion images using an infoGAN

Generative Adversarial Networks (GANs) are a type of generative model that attempts to synthesize novel data that is indistinguishable from the training data [25]. They consist of two neural networks, locked in competition: a generator that captures the data distribution and creates synthesized data (e.g. an image), and a discriminator that estimates the probability that a sample came from the training data rather than from the generator. The two networks are sealed in a zero-sum game, where the success of one corresponds to a failure of the other. The training procedure for the generator is to maximize the probability of the discriminator making an error [26]. Thus, this framework is based on a value function that one model seeks to maximize and the other seeks to minimize. Since the two networks are differentiable, the system generates a gradient that can be used to steer both networks to the right direction.

We have trained an InfoGAN (Information Maximizing Generative Adversarial Network) composed of a Generator and a Discriminator, on all high resolution images of the HAM10000 dataset [27]. The InfoGAN was adapted to the progressive architecture of the model by splitting the structured code to parts and feeding each part to the network by conditioning activation in the corresponding block of the Generator (Supplementary Figure 1). To avoid detrimental competition between Discriminator and Generator, and to achieve convergence in an efficient way, we have followed the recommendations detailed on Chen et al. [27]. Briefly, Discriminator and Generator were composed in progressive 8 blocks with an input/output of spatial resolution of 4×4 in the initial step up to 512 x512 in the step 7. The Batch size have gone dynamically from 128 in step 0 to 2 in the last step. Both, Generator and Discriminator were optimized using Adam with an initial learning rate of 0.0075 and exponential decay of 0.99 and a Wasserstein function with gradient penalty. The training has progressed in phases of progressive increased resolution. More specifically, the model was capable of generating high resolution images with isolated semantic features controlled by a set of real valued variables. Colour, age, sex, localization and type of lesion were the most important semantic features discovered during training in a completely unsupervised fashion without human input. After training, the Generator have produced novel images, similar to those in the dataset.

### 2.5 Contests among general practitioners

To compare the accuracy of DNNs with non-dermatologist practitioners, we conducted two different challenges. The first has been aimed to establish the accuracy of general practitioners in classifying images from the HAM10000 dataset without time constraint. For this purpose, a group of 22 general practitioners from any given center in Buenos Aires (Argentina) were given access to 163 images of the different skin pigmented lesions through an anonymous website specifically created for this purpose. Physicians could enter and exit the website without limitation. Alongside the image, recorded factors such as age, sex and localization of the lesion were shown. All physicians were asked to classify every image among onte within the seven different diagnostics. No incentives were offered for participation. To ensure fair comparisons between the results determined by general practitioners and those determined by the DNNs, the same 162 images were run with the DNN framework which had the worse accuracy metrics in low-resolution and without aggregated clinical features of the eight DNNs tested.

To determine if physicians could benefit from access to the algorithmic tool during dthe own classification task, a second evaluation was conducted. A group of 19 general practitioners that voluntarily accepted to participate in the study was first asked to assess 35 images in a simulated exercise with time constraints (physicians had 45 seconds to classify every image). In a second step, physicians had access to the predictions of the same algorithmic framework used during the first challenge, the same group having classified each image based on both, their criteria and the algorithmic output. For this task, a new set of 35 images was shown with the same time constraints. In both contests the ethics committee waived ethical approval owing to the use of anonymized dermatologic images obtained from the publicly available HAM10000 dataset.

### 2.6 Statistical analysis

After the model had been trained, a test step was performed in which 1,702 images of the seven dermatological manifestations has been used as input and the results were statistically analyzed. A confusion matrix was constructed based on comparing the frameworks’ prediction with each of the actual labels. All analyses were performed and programmed via a Jupyter notebook in Python. Sensitivity, specificity, geometric mean, accuracy and error rate were calculated for each dermatological manifestation [28]. Sensitivity or true positive rate (TPR) represented the positive and correctly classified samples to the total number of positive samples. Specificity or true negative rate (TNR) was estimated as the ratio of the correctly classified negative samples to the total number of negative samples. Geometric means was calculated by using the product of TPR and TNR. Accuracy was defined as the ratio between the correctly classified samples to the total number of samples [28]. We have also calculated the error rate as the complement of accuracy. All these measures are suitable to evaluate the classification performance based on imbalanced data as found in the HAM10000 database. All metric results were calculated with respect to the class labels documented in the HAM10000 database archive.

## 3. Results

### 3.1 Classification metrics across eight different DNNs

The results of global accuracy and error rate for each DNN for the classification of seven pigmented skin lesions at low image resolution (300 × 224 RGB) are shown in Table 2. The average global accuracy for the 8 DNN reached 76.30%±2.79, ranging from 74.05% (EfficientNetB5) to 82.47% (MobileNet). As shown in Supplementary Table 1, TPR, TNR and geometric mean for each disease subtype varied according to the tested DNN. Almost all DNNs showed the highest TPR for melanocytic nevi classification when compared to the other pigmented lesions, with observed geometric mean values equal or lower than 0.65; interestingly, VGG16, VGG19 and MobileNet also showed high TPR for vascular lesion classification (geometric mean values lower than 0.65). In the case of melanoma and benign keratosis classification, all of them showed TPR of approximately 0.5, with geometric mean values around 0.75. Likewise, basal cell carcinoma classification showed TPR of approximately 0.5 and geometric mean values higher than 0.75 using ResNet50, ResNet101, SEResNet50 and EfficientNetB5. Similar results were observed for SEResNet5 and MobileNet and intraepithelial carcinoma classification.

**Table 2.** Classification metrics of HAM10000 images at two different resolutions and without aggregated clinical features using eight different DNNs.

| DNN | Low-resolution images<sup>1</sup> |  | High-resolution images |  |
| --- | --- | --- | --- | --- |
|  | Accuracy<sup>2</sup> | Error rate | Accuracy | Error rate |
| ResNet34 | 75.32 | 24.68 | 76.73 | 23.27 |
| ResNet50 | 74.56 | 25.44 | 75.97 | 24.03 |
| ResNet101 | 75.62 | 24.38 | 77.02 | 22.98 |
| SEResNet50 | 77.82 | 22.22 | 79.13 | 20.87 |
| VGG16 | 76.85 | 23.15 | 78.25 | 21.75 |
| VGG19 | 74.21 | 25.79 | 75.62 | 24.38 |
| EfficientNetB5 | 74.05 | 25.91 | 75.50 | 24.5 |
| MobileNet | 82.47 | 17.53 | 83.88 | 16.12 |

<sup>1</sup> Low-resolution image: 300 x 224 RGB; High-resolution image: 300 x 224 RGB.
<sup>2</sup> Accuracy and error rate are expressed as percentages.

Using higher image resolution (600 × 450 RGB) improved the global accuracies across all DNNs (Table 2). The global accuracy average for the 8 DNNs tested was 77.76±2.77, ranging from 75.50% (EfficientNetB5) to 83.88% (MobileNet). Although higher than the mean average observed with low-resolution images, the difference was not statistically significant (p=0.07; Mann-Whitney U test). The TPR, TNR and geometric mean values for each disease subtype is detailed in Supplementary Table 2. The highest TPR was observed for melanocytic nevi across all DNNs, and in the case of vascular lesions using VGG16 and MobileNet. On the contrary, the lowest TPR was observed for dermatofibrosis and intraepithelial carcinoma with ResNet34, ResNet50, SEResnet50 and EfficientNetB5. When compared with the low-resolution images parallel cases, TPR, TNR and geometric mean values were quite similar for most of the disease subtypes. However, a drastic improvement of TPR was observed for dermatofibrosis using ResNet34 (from 0.23 to 0.34), ResNet50 (from 0.29 to 0.55), ResNet101 (from 0.26 to 0.42), VGG16 (from 0.17 to 0.4), VGG19 (from 0.26 to 0.40) and MobileNet (from 0.39 to 0.5). Similar TPR improvements were observed for intraepithelial carcinoma classification an VGG19(from 0.26 to 0.40). The average cascade framework runtime of the high-resolution classification model was 21.46+/-2.3 milliseconds per image, whereas the low-resolution model required only 18.6+/-1.22 milliseconds per image. Altogether, these results indicate that the tested DNNs can classify seven different types of pigmented skin lesions with accuracies higher than 0.7. No major differences on runtime were observed between the cascade framework input with low- or high resolution.

### 3.2 Classification metrics of different DNNs aggregating image and clinical features

We then investigated if adding clinical features to the analysis could improve the classification accuracy of each DNN. As HAM10000 dataset provides sex, age and localization of the skin lesion associated to every image, we gathered these clinical features and aggregated them with the corresponding image to be used as input for each of the tested DNNs. Results are shown in Table 3. For low-resolution images, the addition of clinical data improved the global performance of all DNNs but MobileNet. The average global accuracy for the 8 DNNs was 78.86%±1.81, ranging from 75.73% (EfficientNetB5) to 81.24% (MobileNet). This represented a statistically significant increase in comparison to global accuracy of DNNs tested with low-resolution images without aggregated clinical features (p=0.004; Mann-Whitney U test). The highest increase was observed for VGG19 raising from 74.21% to 79.43 (5.22%). As shown in Supplementary Table 3, classification improvements were observed in almost all pigmented skin lesions for all DNNs. The highest marginal increases in accuracy were observed for dermatofibrosis where TPR values for ResNet 50, SEResNet50, VGG16, EfficientNetB5 and MobileNet raised 15%, 51%, 22%, 28% and 46%, respectively. Similarly, in the case of intraepithelial carcinoma condition, TPR values increased 13% and 26% for SEResnet50 and VGG19, respectively, and for basal cell carcinoma classification, increased 11%, 14% and 12% for ResNet50, VGG19 and EfficientNetB5, respectively. For vascular lesions, TPR values increased 34%, 14% and 29% for ResNet101, VGG19 and EfficientNetB5, respectively. Finally, adding clinical features to skin images also improved melanoma condition accuracy for VGG16 (from 0.47 to 0.61) and VGG19 (from 0.44 to 0.58). Of note, a decreased of TPR was observed for ResNet101 in classification of dermatofibrosis (from 0.26 to 0.16) and intraepithelial carcinoma (from 0.37 to 0.29), and in melanoma for MobileNet (from 0.90 to 0.72).

**Table 3.** Classification metrics of HAM10000 low- and high-resolution images with aggregated clinical data using eight different DNNs.

| DNN | Low-resolution images<sup>1</sup> |  | High-resolution images |  |
| --- | --- | --- | --- | --- |
|  | Accuracy<sup>2</sup> | Error rate | Accuracy | Error rate |
| ResNet34 | 77.43 | 22.57 | 78.84 | 21.16 |
| ResNet50 | 79.31 | 20.69 | 80.72 | 19.28 |
| ResNet101 | 77.55 | 22.45 | 78.96 | 21.04 |
| SEResNet50 | 80.01 | 19.99 | 80.72 | 19.28 |
| VGG16 | 80.25 | 19.75 | 81.65 | 18.35 |
| VGG19 | 79.43 | 20.57 | 79.02 | 20.98 |
| EfficientNetB5 | 75.73 | 24.27 | 77.14 | 22.86 |
| MobileNet | 81.24 | 18.76 | 84.73 | 15.27 |

<sup>1</sup> Low-resolution image: 300 x 224 RGB; High-resolution image: 300 x 224 RGB.
<sup>2</sup> Accuracy and error rate are expressed as percentages.

For high-resolution images, the average global accuracy also increased when clinical features were added to the model (Table 3). The average global accuracy was 80.22%±2.30, ranging from 77.14% (EfficientNetB5) to 84.73% (MobileNet). This represented a statistically significant increase in comparison to global accuracy of DNNs tested with high-resolution images without aggregated clinical features (p=0.02; Mann-Whitney U test). This was particularly evident in the case of ResNet50 performance with an increase of 4.75%. TPR, TNR and geometric mean values are shown in Supplementary Table 4. Major TPR improvements were observed for dermatofibrosis with ResNet34 (26%), ResNet101 (20%), VGG19 (17%) and EfficientNetB5 (28%). TPR increases were also observed for basal cell carcinoma with ResNet50 (12%) and EfficientNetB5 (13%), for vascular lesions with ResNet101 (37%) and EfficientNetB5 (28%), for melanoma with VGG16 (14%), VGG19 (32%) and MobileNet (16%) and intraepithelial carcinoma with VGG16 (24%). Interestingly, TPR of ResNet50 and SEResNet50 were reduced for dermatofibrosis from 0.55 to 0.20 and from 0.44 to 0.29, respectively. When compared to the DNNs performance with low-resolution images and clinical features no major differences were observed (p=0.21; Mann-Whitney U test). Of interest, for dermatofibrosis classification, TPR increased for ResNet34 (35%), ResNet101 (50%), VGG16 (50%) and VGG19 (24%); the exception was ResNet50 with a TPR reduction from 0.44 to 0.2. Altogether, these results indicate that the addition of information related to sex, age and localization of the lesion improves the accuracy of DNNs.

**Table 4.** Classification metrics of nondermatologists, general practitioners with or without use of the algorithmic platform and with time constraints.

| DNN | Accuracy<sup>1</sup> | Error rate |
| --- | --- | --- |
| EfficientNetB5 | 77.14 | 22.86 |
| GPs<sup>2</sup> | 17.29 | 82.71 |
| GPs + AI | 42.43 | 57.57 |

<sup>1</sup> Accuracy and error rate are expressed as percentages
<sup>2</sup> GPs: general practitioners; AI: artificial intelligence

### 3.4 Performance across synthetic pigmented-skin lesion images

We have applied an infoGAN to generate synthetic images from the HAM10000 dataset. As shown in Figure 2, the synthetic images seemed realistic and diverse. We have then calculated global accuracy for EfficientNetB5 for the classification of seven pigmented skin lesions. Of the 40 synthetic images analyzed, the network made a single error, so the certainty index was 97.5%; however, this value lacks any relevance since the GAN used for pseudo-labelling deliberately increases the definition limit of each class, inducing an improvement in the certainty of the classifier. Altogether, these data indicate that the synthetic samples are highly realistic and can be used as inputs to train DNNs on pigmented skin lesion classification.

**Figure 2.**
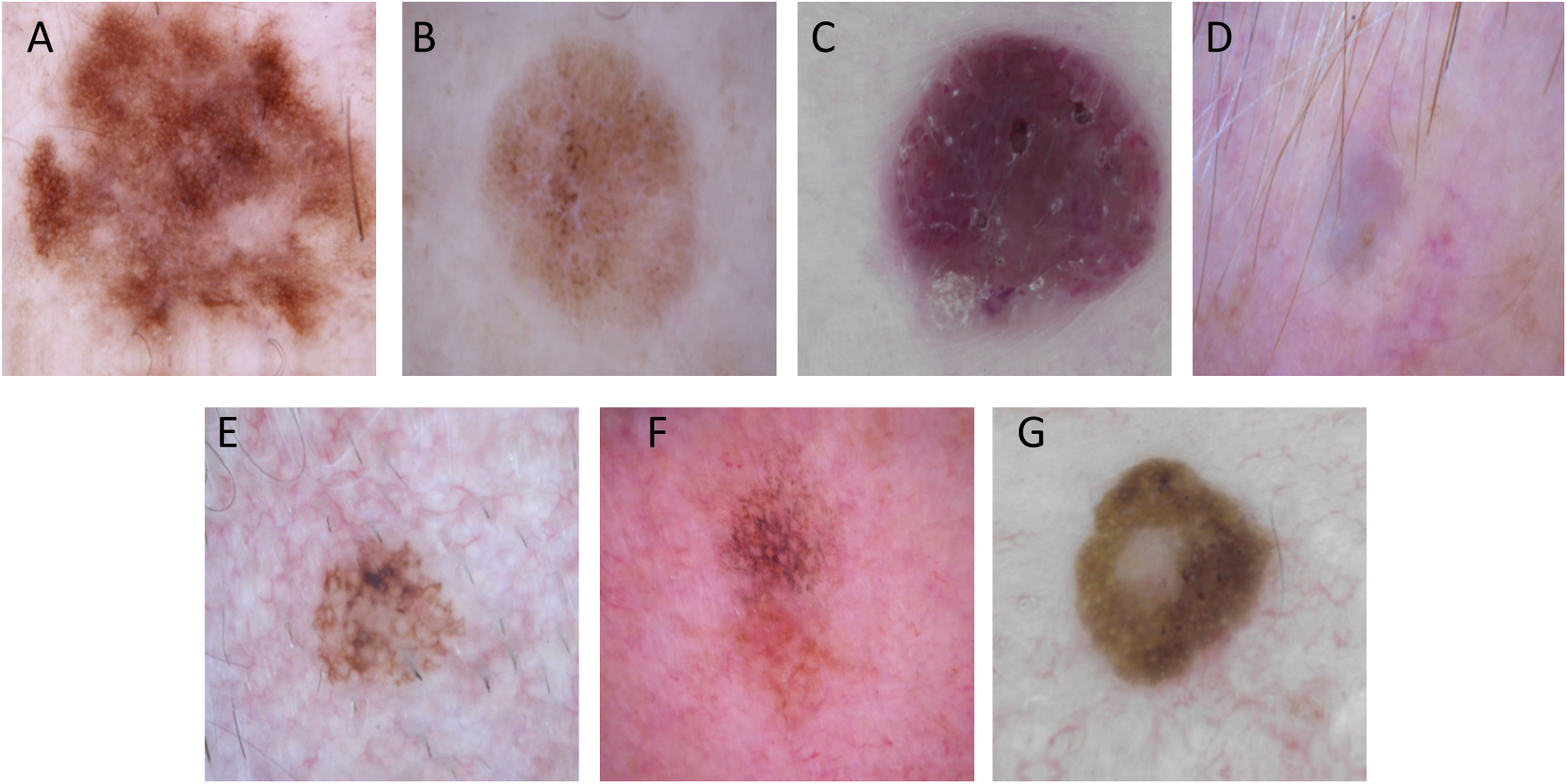
Examples of synthetic images generated with the infoGAN. (A) Melanocytic nevus; (B) Bening keratosis; (C) Vascular lesion; (D) Dermatofibroma; (E) Intraepithelial carcinoma; (F) Basal cell carcinoma; (G) Melanoma. Legends inside each image represents clinical data such as age, sex and localization associated to the image. F: female; M: male; LE: lower extremity; B: back; H: Hand; T: trunk.

### 3.3 Performance across general practitioners with and without assistance from DNNs output

In the first challenge, 22 general practitioners were asked to classify 162 images without any time constraint. The mean global accuracy and mean error rate were 27.74% and 72.26%, respectively. These results were similar to previously published for non-dermatologists [9, 10]. The best TPR (0.79) was obtained for the melanocytic nevi while the worse metrics were observed for vascular lesions (0.02), dermatofibrosis (0.01) and intraepithelial carcinoma (0.07) (Supplementary Table 5). As EfficientNetB5 was slightly less accurate than the other tested DNNs, we have decided to use this framework as comparator (see Table 1). In the same dataset, this DNN had a mean global accuracy of 78.40% and mean error rate 21.60% (Table 4). Compared to physicians, this was a relevant and significant difference. As EfficientNetB5 showed higher TPR in all disease subtypes (Supplementary Table 5).

In the second challenge, 19 general practitioners were asked to classify 35 images with a time constraint of 45 seconds per image. The global accuracy for this classification was 17.29% (Table 4). In the same dataset, EfficientNetB5 achieved a global accuracy of 77.14%, significantly outperforming physicians.

When general practitioners were given the opportunity to access the output of EfficientNetB5 per image, global accuracy raised to 42.42%. This represented an increase of 25.13%. This result also indicated that, in some cases, physicians did not follow the recommendation of the DNN. Of note, access of physicians to DNN prediction increased TPR for basal cell carcinoma (from 0.10 to 0.54) and melanoma (from 0.08 to 0.35) (Supplementary Table 6). In contrast, a small decreased in TPR for benign keratosis was observed (see Supplementary Table 6).

Altogether, these results show that DNNs have the capability to classify seven different pigmented skin lesions with a level of competence higher to that of the general practitioners participating in these challenges. The access of DNN output by physicians improves their ability to classify pigmented skin lesions, particularly basal cell carcinoma and melanoma.

## 4. Discussion

Our results demonstrate that deep learning frameworks trained on large, open-source image datasets can help non-dermatologist physicians in improving their accuracy to categorize the seven most frequent pigmented skin lesions. Additionally, we show that image resolution does not affect the performance of 8 different DNNs. Instead, the aggregation of clinical features (age, sex and lesion localization) significantly increase DNNs performance in both with low-resolution and with high-resolution image inputs. The use of artificial intelligence as a diagnostic aid is a growing trend in dermatology. A digital automated skin assistance tool provides an undeniable help for dermatologists and general practitioners to reduce the morbidity and mortality linked to dermatological diseases by favoring early diagnosis and by the avoidance of unnecessary procedure. The advent of deep/machine learning algorithms has made automated classification of cutaneous lesions an achievable target milestone [29].

Different dermatologic studies have reported early success in the classification of pigmented skin lesions from both clinical and dermoscopic images with a level of accuracy comparable to that of dermatologists. Esteva et al were among of the first one to describe a DNN that performed as well as dermatologists when identifying images with malignant lesions [15]. The authors used a GoogleNet Inception v3 architecture that was pre-trained on approximately 1.28 million images. Then, they have used 129,450 skin images of 2032 different diseases to train and ultimately validated the system using two classes (benign/malignant). The model was compared to the performance of 21 dermatologists using a test set of 135 biopsy-proven lesion clinical and dermoscopic images. The performance of this binary classification method was on par with that of all of the dermatologists who participated. Haenssle et al. presented a very similar approach to Esteva et al [1]. They have compared the diagnostic performance of 58 dermatologists with a GoogLeNet Inception v3 model that was adapted for skin lesion classification with transfer learning, whereby the weights were fine-tuned in all layers. The analysis was limited to dermoscopic images of melanoma vs. benign nevi. In the test dataset of 300 biopsy-proven images, the accuracy of the DNN compared favorably with the one by dermatologists. Likewise, Han et al presented a ResNet152 classifier for 12 different skin diseases based on clinical images that performed comparably to the performance of 16 dermatologists [16]. Fujisawa et al. used a dataset of 4,867 clinical images to train a DNN to differentiate 14 different clinical conditions that included both malignant and benign conditions [30]. The machine’s performance was then compared against that of 13 dermatologists and nine dermatology trainees and tested on 1,142 images distinct to those used for training. The DNN outperformed the dermatologists across every field. Additionally, a set of other recent studies also reached dermatologist-level skin cancer classification by using DNNs [31-34].

In contrast to all these previously mentioned publications comparing the performance across different DNN configurations to the one by dermatologists, our study was carried out with non-dermatologist practitioners. Our results show that the tested frameworks classify pigmented skin lesions much better than the general practitioners that participated in this study. Our results are similar to those published recently by Tschandl et al [35]. These authors have matched a set of DNNs with human readers for the diagnosis of the 7 clinically relevant types of pigmented skin lesions analyzed in our study using the HAM10000 dataset. From the 511 human readers involved in their study, 83 were general practitioners. The authors showed that the top 3 DNNs outperformed physicians with respect to most outcome measures. However, human metrics were only disclosed for dermatology experts, and thus, we cannot compare our metrics to them. Although promising, our results should be analyzed within context as they are derived from a set of pre-existing images and not from a real-life patient observation. Indeed, general practitioners are not trained to diagnose over an image, particularlly if they have just a few seconds to decide. Moreover, in a real-world situation, they would consider other clinical features besides a skin image and given complementary data; they would be evaluating the patient as a whole, not just a skin lesion. In spite of this, our results showing that physician’s access to a DNN output improved their ability to classify pigmented skin lesions are encouraging. Moreover, this assistance improved the positive classification of basal cell carcinomas, one of the most common of all types of cancer, and the most dangerous melanoma.

At a more technical level, our results are in agreement with various other publications that have also demonstrated the capacity of multiple DNN constructs to classify clinical or dermoscopic skin images [17, 36-42]. Some of these classifiers have been translated onto online platforms and smartphone applications for use by dermatologists or individuals in the community setting (e.g. modelderm, MoleMapper, MoleAnalyzer Pro) [43]. Most of these published works use non-public archives [1, 15]. This makes it very difficult to reproduce the results and compare the performance of published classifiers against each other. Thus, we have decided to compare the performance of 8 different DNNs using a unique and public dataset as the HAM10000. Among other things, we show that the quality of the input images marginally affects the performance of a classification task for a given DNN, as similar has been achieved both with input with low- and high-resolution image. According to our results, the higher the resolution of the image, the better the performance of a given DNN is; however, the improvements were not so evidently compared to those observed with the addition of a few clinical features to the analysis. Indeed, algorithms trained with low-resolution images and aggregated clinical features achieve levels of precision similar to those obtained with better resolution images without clinical features. This is in line with a recent publication that showed that adding clinical information to skin lesion images improves the diagnostic accuracy of dermatologists [1]. This result is significant as it implies that adding clinical features is more important than the resolution of the input image, being the best situation the combination of high-resolution images and clinical features. From a clinical perspective, it is important to note that other authors have used other complex mathematical techniques to improve algorithm’s performance, such as dropout, data augmentation and batch normalization [44-46]. Data augmentation along with a larger database including both higher resolution images and clinical data such as symptoms and localization of such image could sensibly improve the image with higher probability.

In this work, we have used the HAM10000 dataset from the ISIC archive (https://isic-archive.com/) [21]. As members of the scientific community, we are truly grateful to the researchers who have generated this dataset and we acknowledge the enormous effort invested on it. Because of permissive licensing, well- structured availability, and large size it is currently the standard source for dermoscopic image analysis research. Although other open skin lesions datasets containing clinical and dermoscopic images are available, these are not as large as the HAM10000, leaving it as the only public one that can be used for training and validation of new algorithms [47]. We have set out to solve this problem by generating synthetic images using an infoGAN. This framework consists of a generator network that try to produce realistically looking images, while a discriminator network aims to classify well between samples coming from a real training data and fake samples generated by the generator [25, 26]. Our results show that the synthetic skin images can be used as input images by DNNs, similar to that observed with real images of the HAM10000 dataset. This indicates that this synthetic dataset can be used to train and test different algorithmic frameworks, overcoming the lack of skin image databases representing the diversity of skin types observed in the real world. As described in Tschandl et al. the HAM10000 dataset presents some flaws [21]. First, it is biased towards melanocytic lesions (12,893 of 13,786 images are nevi or melanomas). Likewise, although it is an excellent curated image dataset, it is composed mostly of skin images of a mostly fair-skinned Caucasian population thus making difficult to extrapolate the results to other racial groups. Indeed, it was recently reported that a DNN trained on an image dataset composed mainly of Caucasian population skin images (Fitzpatrick 1 and 2 skin types) could not be extrapolated to African-black skin color patients [48]. In that study, the DNN’s accuracy was as low as 17%. Other open skin lesions datasets containing clinical and dermoscopic images from non-Caucasian human skin types are available, however these are not as large as the HAM10000 or are not publicly available [47]. To tackle this problem, and based on the results shown in this work, we are generating synthetic images representing the different pigmented lesions of the HAM10000 dataset on the six human skin types according to the Fitzpatrick scale using an infoGAN [27].

In terms of limitations within our study, firstly, the algorithms were bound to only in seven different disease classes which does not reflect clinical reality as many more options should be taken into account when diagnosing [49]. As a consequence, the use of these classification algorithms should be regarded as an assisting tool for dermatologists or general practitioners that may improve accuracy within a limited scope but not as a replacement for independent diagnoses qualified by a supervising physician. Secondly, deep learning models are powerful “black box” models which remain relatively uninterpretable compared to the statistical methods used in medical practice [50]. Computer vision models combine pixel-based visual information in a highly intricate way, making it difficult to link model output back to the visual input. A third limitation is that although the test dataset was disjunct from the training dataset, all the images belonged to the same database (HAM10000), raising concerns about their ability to generalize on a truly external test set coming from a different image bank. It is known that the efficacy of DNNs varies based on the set of images with which they are trained. Each model may have different sensitivities and specificities and may be subject to a unique set of biases and shortcomings in prediction introduced by the image training set. In a recent study, a binary-classification DNN for melanocytic nevus vs. melanoma, trained on ISIC images, showed good performance on an ISIC test dataset but performed badly on an external test dataset from the PH2 dermoscopic image source [1, 47]. Using just 100 images from the external database for fine-tuning the DNN sufficed to completely restore the original performance. Another important issue is related to the artifacts observed in clinical or dermoscopic images, such as surgical skin markings, dark corners, gel bubbles, superimposed color charts, overlayed rulers, and occluding hair that can affect image classification by automated algorithms [51]. Various methods have been reported for the removal of such artifacts and strategies for preprocessing of images were described to improve the classification outcomes of DNNs [51-54].

In conclusion, our findings show that deep learning algorithms can successfully assist non-dermatologist physicians in potentiating their classification performance across seven different pigmented skin lesions. Moreover, this technology would help primary care physicians in the decision-making process on which patients are at highest risk for skin cancer, with subsequent referral to dermatology for total body skin examination. These models could be easily implemented in a mobile app, on a website, or even integrated into an electronic medical record system enabling fast and cheap access skin screenings, even outside the hospital. Future research should carefully validate our results using other image datasets containing patients across a blend of different ages and ethnicities, including additional cutaneous lesions and color skin types. Ultimately, automated diagnostic systems based on DNNs will allow clinicians to enhance patient care by means of improving their classification skills outside of their field of expertise.

## Data Availability

This study used images from the anonymous and annotated HAM10000 dataset publicly available through the International Skin Imaging Collaboration (ISIC) archive

## 5. Acknowledgements

This research did not receive any specific grant from funding agencies in the public, commercial, or not-for-profit sectors.

## 6. Conflict of interest

Maximiliano Lucius and Carlos M. Galmarini are associates at Topazium. Martín Belvisi is an advisor of Topazium. Jorge De All, José Antonio De All, Marisa Lanfranconi and Victoria Lorenzatti are associates at Sanatorio Otamendi. Luciana Radizza is an associate of IOSFA.

## Notes

### Competing Interest Statement

The authors declare the following financial interests/personal relationships which may be considered as potential competing interests: Maximiliano Lucius and Carlos M. Galmarini are employees of Topazium. Jorge De All, José Antonio De All and Victoria Lorenzatti are employees at Sanatorio Otamendi. Luciana Radizza is an employee of IOSFA

### Funding Statement

No external funding was received

## Reference

[1]. Haenssle HA, Fink C, Schneiderbauer R, et al. Man against machine: diagnostic performance of a deep learning convolutional neural network for dermoscopic melanoma recognition in comparison to 58 dermatologists. Ann Oncol 2018; 29: 1836–42.

[2]. Annessi G, Bono R, Sampogna F, et al. Sensitivity, specificity, and diagnostic accuracy of three dermoscopic algorithmic methods in the diagnosis of doubtful melanocytic lesions: the importance of light brown structureless areas in differentiating atypical melanocytic nevi from thin melanomas. J Am Acad Dermatol 2007; 56: 759–67.

[3]. Argenziano G, Soyer HP. Dermoscopy of pigmented skin lesions--a valuable tool for early diagnosis of melanoma. Lancet Oncol 2001; 2: 443–9.

[4]. Brochez L, Verhaeghe E, Grosshans E, et al. Inter-observer variation in the histopathological diagnosis of clinically suspicious pigmented skin lesions. J Pathol 2002; 196: 459–66.

[5]. Russo T, Piccolo V, Lallas A, et al. Dermoscopy of Malignant Skin Tumours: What’s New? Dermatology 2017; 233: 64–73.

[6]. Witkowski AM, Ludzik J, Arginelli F, et al. Improving diagnostic sensitivity of combined dermoscopy and reflectance confocal microscopy imaging through double reader concordance evaluation in telemedicine settings: A retrospective study of 1000 equivocal cases. PLoS One 2017; 12:e0187748.

[7]. Kittler H, Pehamberger H, Wolff K, et al. Diagnostic accuracy of dermoscopy. Lancet Oncol 2002; 3: 159–65.

[8]. Vestergaard ME, Macaskill P, Holt PE, et al. Dermoscopy compared with naked eye examination for the diagnosis of primary melanoma: a meta-analysis of studies performed in a clinical setting. Br J Dermatol 2008; 159: 669–76.

[9]. Federman DG, Concato J, Kirsner RS. Comparison of dermatologic diagnoses by primary care practitioners and dermatologists. A review of the literature. Arch Fam Med 1999; 8: 170–2.

[10]. Federman DG, Kirsner RS. The abilities of primary care physicians in dermatology: implications for quality of care. Am J Manag Care 1997; 3: 1487–92.

[11]. Moreno G, Tran H, Chia AL, et al. Prospective study to assess general practitioners’ dermatological diagnostic skills in a referral setting. Australas J Dermatol 2007; 48: 77–82.

[12]. Suneja T, Smith ED, Chen GJ, et al. Waiting times to see a dermatologist are perceived as too long by dermatologists: implications for the dermatology workforce. Arch Dermatol 2001; 137: 1303–7.

[13]. Tran H, Chen K, Lim AC, et al. Assessing diagnostic skill in dermatology: a comparison between general practitioners and dermatologists. Australas J Dermatol 2005; 46: 230–4.

[14]. Hogarty DT, Su JC, Phan K, et al. Artificial Intelligence in Dermatology-Where We Are and the Way to the Future: A Review. Am J Clin Dermatol 2019;

[15]. Esteva A, Kuprel B, Novoa RA, et al. Dermatologist-level classification of skin cancer with deep neural networks. Nature 2017; 542: 115–8.

[16]. Han SS, Kim MS, Lim W, et al. Classification of the Clinical Images for Benign and Malignant Cutaneous Tumors Using a Deep Learning Algorithm. J Invest Dermatol 2018; 138: 1529–38.

[17]. Marchetti MA, Codella NCF, Dusza SW, et al. Results of the 2016 International Skin Imaging Collaboration International Symposium on Biomedical Imaging challenge: Comparison of the accuracy of computer algorithms to dermatologists for the diagnosis of melanoma from dermoscopic images. J Am Acad Dermatol 2018; 78:270–7 e1.

[18]. Tschandl P, Argenziano G, Razmara M, et al. Diagnostic accuracy of content-based dermatoscopic image retrieval with deep classification features. Br J Dermatol 2019; 181: 155–65.

[19]. Hekler A, Utikal JS, Enk AH, et al. Superior skin cancer classification by the combination of human and artificial intelligence. Eur J Cancer 2019; 120: 114–21.

[20]. Fitzpatrick TB. The validity and practicality of sun-reactive skin types I through VI. Arch Dermatol 1988; 124: 869–71.

[21]. Tschandl P, Rosendahl C, Kittler H. The HAM10000 dataset, a large collection of multi-source dermatoscopic images of common pigmented skin lesions. Sci Data 2018; 5: 180161.

[22]. Simonyan K, Zisserman A. Very deep convolutional networks for large-scale image recognition. arXiv 2014; 1409.556.

[23]. Hu J, Shen L, Albanie S, et al. Squeeze-and-Excitation Networks arXiv 2019; 1709.01507v4.

[24]. Tan M, Le QV. EfficientNet: Rethinking Model Scaling for Convolutional Neural Networks. arXiv 2019; 1905.11946v2.

[25]. Goodfellow I, Pouget-Abadie J, Mirza M, et al. Generative adversarial nets. Neural Information Processing Systems 2014. Montreal, Canada. 2014.

[26]. Goodfellow I, Pouget-Abadie J, Mirza M, et al. Generative adversarial nets. arXiv 2014; 1406.2661.

[27]. Chen X, Duan Y, Houthooft R, et al. Infogan: interpretable representation learning by information maximizing generative adversarial nets. arXiv 2016; 1606.03657.

[28]. Tharwat A. Classification assessment methods. Applied Computing Informatics 2018; 15: 1–13.

[29]. LeCun Y, Bengio Y, Hinton G. Deep learning. Nature 2015; 521: 436–44.

[30]. Fujisawa Y, Otomo Y, Ogata Y, et al. Deep-learning-based, computer-aided classifier developed with a small dataset of clinical images surpasses board-certified dermatologists in skin tumour diagnosis. Br J Dermatol 2018; 180: 373–81.

[31]. Brinker TJ, Hekler A, Enk AH, et al. Deep neural networks are superior to dermatologists in melanoma image classification. Eur J Cancer 2019; 119: 11–7.

[32]. Brinker TJ, Hekler A, Enk AH, et al. A convolutional neural network trained with dermoscopic images performed on par with 145 dermatologists in a clinical melanoma image classification task. Eur J Cancer 2019; 111: 148–54.

[33]. Brinker TJ, Hekler A, Enk AH, et al. Deep learning outperformed 136 of 157 dermatologists in a head-to-head dermoscopic melanoma image classification task. Eur J Cancer 2019; 113: 47–54.

[34]. Brinker TJ, Hekler A, Hauschild A, et al. Comparing artificial intelligence algorithms to 157 German dermatologists: the melanoma classification benchmark. Eur J Cancer 2019; 111: 30–7.

[35]. Tschandl P, Codella N, Akay BN, et al. Comparison of the accuracy of human readers versus machine-learning algorithms for pigmented skin lesion classification: an open, web-based, international, diagnostic study. Lancet Oncol 2019; 20: 938–47.

[36]. Nasr-Esfahani E, Samavi S, Karimi N, et al. Melanoma detection by analysis of clinical images using convolutional neural network. Conf Proc IEEE Eng Med Biol Soc 2016; 2016: 1373–6.

[37]. Yu C, Yang S, Kim W, et al. Acral melanoma detection using a convolutional neural network for dermoscopy images. PLoS One 2018; 13:e0193321.

[38]. Pomponiu V, Nejati H, Cheung NM. Deepmole: Deep neural networks for skin mole lesion classification. Proceedings of the 2016 IEEE International Conference on Image Processing (ICIP). 2016 IEEE International Conference on Image Processing (ICIP). Phoenix, AZ, USA. 2016.

[39]. Codella N, Cai J, Abedini M, et al. Deep learning, sparse coding, and SVM for melanoma recognition in dermoscopy images. Proceedings of the 6th International Workshop on Machine Learning in Medical Imaging.. 6th International Workshop on Machine Learning in Medical Imaging. Munich, Germany. 2015.

[40]. Kawahara J, BenTaieb A, Hamarneh G. Deep features to classify skin lesions. Proceedings of the 2016 IEEE 13th International Symposium on Biomedical Imaging (ISBI). 2016 IEEE 13th International Symposium on Biomedical Imaging (ISBI). Prague, Czech Republic. 2016.

[41]. Bi L, Kim J, Ahn E, et al. Automatic skin lesion analysis using large-scale dermoscopy images and deep residual networks. arXiv 2017; 1703.04197

[42]. Sun X, Yang J, Sun M, et al. A benchmark for automatic visual classification of clinical skin disease images. Proceedings of the European Conference on Computer Vision. European Conference on Computer Vision. Amsterdam, The Netherlands. 2016.

[43]. Chuchu N, Takwoingi Y, Dinnes J, et al. Smartphone applications for triaging adults with skin lesions that are suspicious for melanoma. Cochrane Database Syst Rev 2018; 12:CD013192.

[44]. Srivastava N, Hinton GE, Krizhevsky A, et al. Dropout: a simple way to prevent neural networks from overfitting. J Mach Learn Res 2014; 15: 1929–58.

[45]. Ioffe S, Szegedy C. Batch normalization: accelerating deep network training by reducing internal covariate shift. arXiv 2015; 1502.03167.

[46]. Lemley J, Bazrafkan S, Corcoran P. Smart augmentation learning an optimal data augmentation strategy. arXiv 2017; 1703.08383.

[47]. Mendonca T, Ferreira PM, Marques JS, et al. PH(2) - a dermoscopic image database for research and benchmarking. Conf Proc IEEE Eng Med Biol Soc 2013; 2013: 5437–40.

[48]. Kamulegeya LH, Okello M, Bwanika JM, et al. Using artificial intelligence on dermatology conditions in Uganda: A case for diversity in training data sets for machine learning.. BioRxiv 2019; 826057

[49]. Maron RC, Weichenthal M, Utikal JS, et al. Systematic outperformance of 112 dermatologists in multiclass skin cancer image classification by convolutional neural networks. Eur J Cancer 2019; 119: 57–65.

[50]. Hulstaert E, Hulstaert L. Artificial intelligence in dermato-oncology: a joint clinical and data science perspective. Int J Dermatol 2019; 58: 989–90.

[51]. Winkler JK, Fink C, Toberer F, et al. Association Between Surgical Skin Markings in Dermoscopic Images and Diagnostic Performance of a Deep Learning Convolutional Neural Network for Melanoma Recognition. JAMA Dermatol 2019; 155: 1135–41.

[52]. Yoshida T, Celebi ME, Schaefer G, et al. Simple and effective pre-processing for automated melanoma discrimination based on cytological findings. IEEE International Conference on Big Data. Washington, DC. USA. 2016.

[53]. Jafari M, Karimi N, Nasr-Esfahani E. Skin lesion segmentation in clinical images using deep learning. 23rd International Conference on Pattern Recognition (ICPR);. Cancun, Mexico. 2016.

[54]. Salido J, Ruiz C. Using deep learning to detect melanoma in dermoscopy images. Int J Mach Learn Comput 2018; 8: 61–8.

